# Detection of mosaic chromosomal alterations in children with severe developmental disorders recruited to the DDD study

**DOI:** 10.1101/2022.03.28.22273024

**Authors:** Ruth Y. Eberhardt, Caroline F. Wright, David R. FitzPatrick, Matthew E. Hurles, Helen V. Firth

**Author notes:** Corresponding author: Helen V. Firth.

## Abstract

**Purpose:** Structural mosaicism has been previously implicated in developmental disorders. We aim to identify rare mosaic chromosomal alterations (MCAs) in probands with severe undiagnosed developmental disorders.

**Methods:** We identified MCAs in SNP array data from 12,530 probands in the Deciphering Developmental Disorders (DDD) study using MoChA.

**Results:** We found 61 MCAs in 57 probands, many of these were tissue specific. In 23/26 (88.5%) cases for which the MCA was detected in saliva where blood was also available for analysis, the MCA could not be detected in blood. The MCAs included 20 polysomies, comprising either one arm of a chromosome or a whole chromosome, for which we were able to show the timing of the error (25% mitosis, 40% meiosis I, 35% meiosis II). Only 2/57 (3.5%) of the probands in whom we found MCAs had another likely genetic diagnosis identified by whole exome sequencing, despite an overall diagnostic yield of ∼40% across the cohort.

**Conclusion:** Our results show that identification of MCAs provides candidate diagnoses for previously undiagnosed patients with developmental disorders, potentially explaining ∼0.45% of cases in the DDD study. Nearly 90% of these MCAs would have remained undetected by analysing DNA from blood and no other tissue.

## INTRODUCTION

Genetic mosaicism is the presence of two or more genetically distinct lineages of cells in one individual, arising from post-zygotic mutations. Mosaic variation can consist of single nucleotide variants (SNVs) and indels or it may involve larger stretches of the genome, including copy number variants (CNVs) and aneuploidies. Mosaicism has been associated with diseases including neurodevelopmental disorders^1-4^. The clinical consequences of mosaicism vary according to the nature of the mosaic event, the stage of development at which this event occurs, and the tissue types in which this event is present^5^.

Very large chromosomal abnormalities, such as complete autosomal aneuploidy, are generally incompatible with life, with the exception of trisomy 21. However, mosaic aneuploidies are better tolerated and have been identified in many autosomes including chromosomes 7, 8, 9, 14, 16, 17, 19 and 22^6-8^. Moreover, children with mosaic trisomies of chromosomes 13 and 18 live for much longer than those with constitutive trisomies, with 80% and 70% of patients with mosaic trisomy 13 and 18 respectively surviving for at least a year compared to 8% with non-mosaic trisomies^9,10^. Additionally, mosaic uniparental disomy (UPD, where two copies of one chromosome are inherited from one parent) has also been associated with several developmental disorders^5,11-14^.

Mosaic chromosomal alterations (MCAs) can be detected in SNP genotyping array data by identifying differences from the expected log R ratio (LRR) and B-allele frequency (BAF). LRR gives a measure of the intensity at any given position on the array and deviations from the expected LRR indicate an abnormal copy number. BAF is a normalized measure of the intensity ratio of two alleles (A and B), such that a BAF of 1 or 0 indicates the complete absence of one of the two alleles (e.g., homozygous AA or BB), and a BAF of 0.5 indicates the equal presence of both alleles (e.g., heterozygous AB). Deviations in BAF can indicate the presence of CNVs or UPD.

There are several tools which use LRR and BAF to detect mosaic chromosomal alterations from SNP array data. Mosaic Alteration Detection (MAD) uses the Genome Alteration Detection Algorithm (GADA) to detect mosaic CNVs and UPDs^15,16^. Parent-of-Origin-based Detection in trios (triPOD) uses an overlapping window approach to detect mosaic CNVs and UPDs in parent-offspring trios, but the absence of parental data makes this tool unsuitable for many cohorts^17^. MONTAGE is a recently developed tool using a sliding window approach for rapid detection of mosaic CNVs, however it is unable to detect mosaic UPDs^18^. MoChA is a bcftools plugin which identifies mosaic CNVs and UPDs in array data using a hidden Markov model (HMM) to detect imbalances in phased BAF and LRR^19,20^. We chose to use MoChA because it is quick to run, sensitive, doesn’t require trio information and is able to detect both mosaic CNVs and UPDs.

The Deciphering Developmental Disorders study (DDD) is a cohort of 13,612 children with severe developmental disorders in the UK and Ireland^21^. The DDD study recruited patients from 2011-2015 who remained undiagnosed following expert review by a clinical geneticist and completion of routine genetic testing. MCAs have previously been investigated in this cohort using whole exome sequencing (WES) from 4,911 probands using MrMosaic and additionally from SNP genotyping array data for 1,303 of these probands using MAD and triPOD^3,4^. However, the majority of DDD probands have not been analysed systematically for the presence of MCAs. Here we used MoChA to detect MCAs across all 12,530 probands with SNP genotyping array data in the DDD study.

## MATERIALS AND METHODS

### Patient cohort

A total of 13,612 probands with developmental disorders, and their parents, were recruited to the DDD study by all 24 Regional Genetics Services in the UK and Republic of Ireland.

Blood-extracted DNA and/or saliva samples were collected from all probands, and where possible saliva samples were collected from parents. SNP genotyping array data was generated for 12,530 of the probands in this study. Probands were systematically phenotyped by consultant clinical geneticists using the Human Phenotype Ontology (HPO)^22^ and a structured questionnaire in DECIPHER (www.deciphergenomics.org)^23^.

### Mosaic chromosomal alteration detection from array

Samples from 1,465 probands were genotyped on the Illumina HumanOmniExpress chip, and samples from 11,065 probands were genotyped on the Illumina HumanCoreExome chip. Intensity data was converted into VCF format including BAF and LRR using gtc2vcf^24^.

Mosaic chromosomal alterations were detected using MoChA^19,20^. The output was filtered to remove samples with: BAF phase concordance across phased heterozygous sites underlying the call of >0.51, calls <100kbp, calls with a LOD score of <10 for the model based on BAF and genotype phase, calls flagged by MoChA as likely germline CNVs, and calls with an estimated cell fraction of >50%. More stringent filters were subsequently applied to identify rare MCAs of likely clinical significance: events which occur in more than 1% of the cohort were removed, events overlapping CNVs previously identified in the cohort were removed, and events which were <1Mb in length were removed unless they overlapped genes known to cause developmental disorders (https://www.ebi.ac.uk/gene2phenotype)^25^. All MCAs remaining after these filters were manually reviewed to evaluate data quality; events with low deviation in BAF, events in regions where the genotyping array had sparse SNPs and events in samples with noisy data were removed.

## RESULTS

### Potentially clinically relevant MCAs were identified in 57 probands with developmental disorders

A total of 28,864 candidate MCAs were identified by MoChA in the 12,530 probands studied. After initial filtering (Methods), 558 candidate MCAs remained. These events comprised 249 duplications, 78 deletions, 109 copy number neutral loss of heterozygosity events and 122 events where the type could not be determined. After further filtering to identify events of potential clinical relevance, 330 events were reviewed manually to evaluate data quality, and subsequently 61 events from 57 probands were identified for clinical evaluation (Figure 1, Supplementary table 1). These MCAs represent a potential diagnostic yield of 0.45% in our cohort. These 61 events comprise 33 duplications, 12 deletions, 14 copy number neutral loss of heterozygosity events, a deletion flanked by copy number neutral loss of heterozygosity and a duplication followed by uniparental disomy of the majority of the q-arm of chromosome 1. The 33 duplications affect 18 different chromosomes, with the most frequently affected being chromosome 12 (six events); the 12 deletions affect seven different chromosomes, of which the most frequently affected is chromosome X (four events); and the UPDs affect nine different chromosomes, of which the most frequently affected is chromosome 13 (four events) (Figure 2). All of these 57 probands had previously undergone WES, but likely diagnostic variants were only identified in two individuals^26^.

**Table 1.** Pathogenic and likely pathogenic MCAs in DDD patients.

| Chr. | Event type<sup>a</sup> | Start-end (GRCh37)<sup>b</sup> | Size (Mb) | Blood %<sup>c</sup> | Saliva %<sup>c</sup> | Syndrome |
| --- | --- | --- | --- | --- | --- | --- |
| 1 | dup+upd | 165589535-249250621 (q) | 83.66 | nd | 39 | Mosaic likely pathogenic duplication - ACMG score 0.90 (G1A, G3C) |
| 2 | del | 223873590-232804522 | 8.93 | 0 | 22 | Mosaic pathogenic deletion - ACMG score 1.90 (L1A, L2A, L3C) |
| 3 | dup | 153567441-198022430 | 44.46 | 0 | 54 | Mosaic pathogenic partial trisomy 3q23-ter |
| 5 | dup | 1-46174864 | 46.18 | nd | 36 | Mosaic trisomy 5p |
| 5 | dup | 123851734-148651711 | 24.80 | 0 | 34 | Mosaic likely pathogenic duplication - ACMG score 0.90 (G1A, G3C) |
| 7 | del | 1-159138663 (w) | 159.14 | 58 | nd | MIRAGE syndrome due to SAMD9 variant |
| 7 | upd | 64864800-159138663 (q) | 94.27 | 7 | 0 | MIRAGE syndrome due to SAMD9 variant |
| 7 | dup | 99227172-159138663 | 59.91 | 0 | 43 | Mosaic pathogenic partial trisomy 7q21.11-ter |
| 8 | dup | 1-146364022 (w) | 146.36 | 45 | nd | Mosaic trisomy 8 |
| 8 | dup | 22487087-29344462 | 6.85 | 25 | nd | Mosaic pathogenic duplication - ACMG score 1.35 (G1A, G2H, G3C, G5A) |
| 8 | del | 101011612-120215645 | 19.20 | nd | 33 | Langer Gideon syndrome (includes EXT1) |
| 9 | dup | 1-141213431 (w) | 141.21 | 19 | nd | Mosaic trisomy 9 |
| 10 | del | 121375181-135534747 | 14.16 | 0 | 43 | Mosaic pathogenic deletion - ACMG score 1.35 (L1A, L2C, L5A) |
| 11 | dup | 41887927-47877800 | 5.99 | 0 | 22 | Mosaic pathogenic duplication - ACMG score 1.35 (G1A G3C, G5A) |
| 12 | dup | 1-26749137 (p) | 26.75 | 0 | 33 | Pallister-Killian syndrome |
| 12 | dup | 1-34523378 (p) | 34.52 | 0 | 34 | Pallister-Killian syndrome |
| 12 | dup | 1-34758266 (p) | 34.76 | 0 | 63 | Pallister-Killian syndrome |
| 12 | dup | 1-34781187 (p) | 34.78 | 0 | 44 | Pallister-Killian syndrome |
| 12 | dup | 1-34801271 (p) | 34.80 | 0 | 25 | Pallister-Killian syndrome |
| 12 | dup | 1-34826574 (p) | 34.83 | 0 | 63 | Pallister-Killian syndrome |
| 13 | dup | 1-115169878 (w) | 115.17 | 0 | 11 | Mosaic trisomy 13 |
| 13 | dup | 1-115169878 (w) | 115.17 | 16 | nd | Mosaic trisomy 13 |
| 13 | del flanked by loh | 33986219-115169878 | 81.18 | 23 | nd | Mosaic pathogenic deletion - ACMG score 2.35 (L1A, L2A, L3C, L5A) |
| 14 | dup | 1-107349540 (w) | 107.35 | nd | 45 | Mosaic trisomy 14 |
| 14 | upd (pat) | 1-107349540 (w) | 85.62 | nd | 36 | Mosaic Kagami-Ogata syndrome |
| 16 | dup | 27183151-34747045 | 7.56 | 0 | 31 | Mosaic pathogenic duplication - ACMG score 1.35 (G1A, G2H, G3C, G5A) |
| 18 | dup | 1-14084928 (p) | 14.09 | nd | 16 | Mosaic pathogenic duplication - ACMG score 1.35 (G1A, G2H, G3C, G5A) |
| 18 | dup | 1-14988113 | 14.99 | 0 | 25 | Mosaic pathogenic duplication - ACMG score 1.35 (G1A, G2H, G3C, G5A) |
| 18 | dup | 1-78077248 (p) | 78.08 | 14 | nd | Mosaic partial trisomy 18 |
| 18 | del | 48368256-78077248 | 29.71 | 0 | 45 | Mosaic partial monosomy 18q12.3-ter |
| 18 | del | 49315539-78077248 | 28.76 | 0 | 49 | Mosaic partial monosomy 18q12.3-ter |
| 19 | dup | 31740744-41525952 | 9.79 | nd | 29 | Mosaic pathogenic duplication - ACMG score 1.35 (G1A, G2H, G3C, G5A) |
| 20 | dup | 1-63025520 (w) | 63.03 | 0 | 22 | Mosaic trisomy 20 |
| 21 | dup | 1-48129895 (w) | 48.13 | nd | 12 | Mosaic trisomy 21 |
| 21 | dup | 1-48129895 (w) | 48.13 | nd | 40 | Mosaic trisomy 21 |
| 21 | dup | 1-48129895 (w) | 48.13 | nd | 30 | Mosaic trisomy 21 |
| 22 | del | 45311891-51304566 | 5.99 | 32 | 53 | Mosaic pathogenic deletion - ACMG score 1.45 (L1A, L2A, L3A, L5A) |
| X | dup | 1-155270560 (w) | 155.27 | 3 | nd | Mosaic XXXY/XXXXY syndrome |
| X | dup | 38541235-41749282 | 3.21 | 19 | nd | Mosaic triple X syndrome |
| X | dup | 38557085-41742515 | 3.19 | 15 | nd | Mosaic triple X syndrome |
| X | del | 1-155270560 (w) | 155.27 | 0 | 43 | Mosaic Turner syndrome |
| X | del | 1-155270560 (w) | 155.27 | nd | 55(p)<br>24(q) | Mosaic Turner syndrome |
| X | del | 1-155270560 (w) | 155.27 | 25 | nd | Mosaic Turner syndrome |
| X | del | 1-155270560 (w) | 155.27 | nd | 28 | Mosaic Turner syndrome |

<sup>a</sup>dup duplication, del deletion, upd uniparental disomy, upd (pat) paternal uniparental disomy
<sup>b</sup>w whole chromosome, p p-arm, q q-arm
<sup>c</sup>nd not done

**Figure 1.**
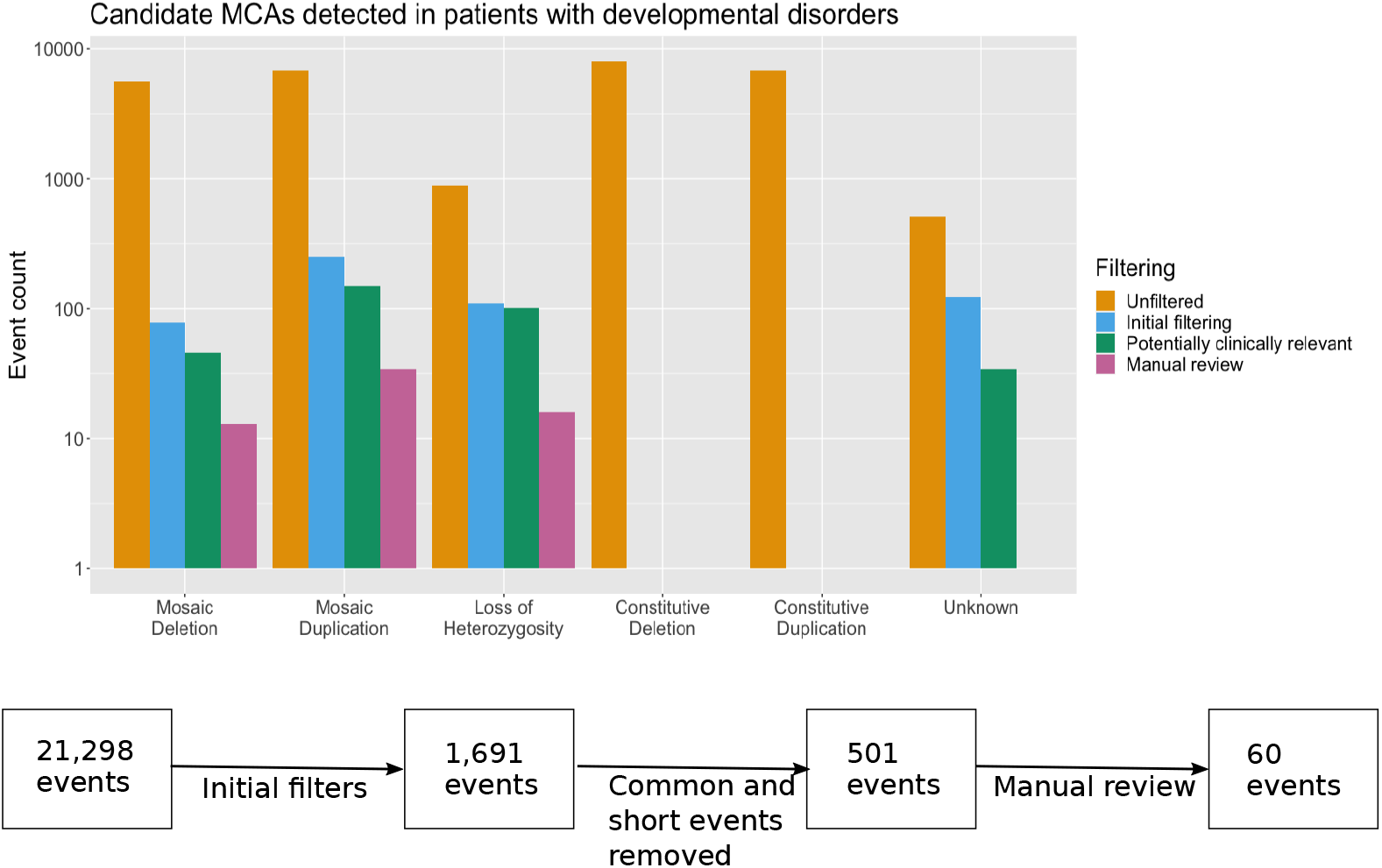
Workflow used to identify potentially clinically relevant MCAs. The flowchart shows the different filtering stages and the total number of events remaining at each stage. The bar plot shows the number of events of each type remaining at each stage.

**Figure 2.**
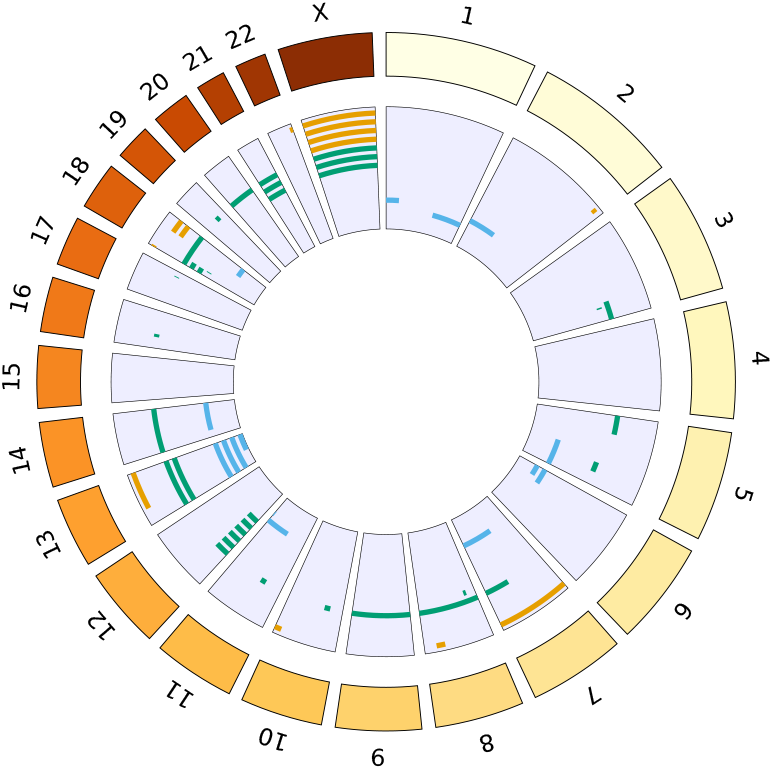
The distribution of MCAs in the genomed. Each bar represents one event; deletions are shown in orange, duplications in green and loss of heterozygosity in blue.

### Tissue specificity was observed for the majority of clinically relevant MCAs

A total of 42 MCAs were detected in saliva from 38 probands (Supplementary table 1). For nine of the probands where a total of 11 MCAs were found in saliva, we also had genotyping array data from blood; the MCA was also detected in only three of these. In an additional 14 probands where15 MCAs were detected in saliva, we also had WES and/or array CGH (aCGH) data from blood; there was *no* evidence of the MCA in any of these. For the remaining 16 MCAs from 15 probands, no other tissue type was available for testing. A total of 22 MCAs were detected in blood from 22 probands. There was SNP array data available from saliva in three of these, and in all three cases the MCA was also detected in saliva. In one additional case, although WES data was available from saliva, this MCA was a mosaic UPD and we are currently unable to detect events of this nature in WES data. For the remaining 18 MCAs detected in blood, no other tissue was available. In the three cases where the MCAs were detected in both saliva and blood, the cell fractions differed by up to 2.3-fold depending on the tissue tested. The mosaic deletion in ID 259003 and the mosaic UPD in ID 274396 were both detected in higher levels in saliva than in blood (ID 259003: saliva 53% blood 32%, ID 274396: saliva 41%, blood 18%), in contrast the mosaic loss-of-heterozygosity in ID 257978 is observed at a higher level in blood (30%) than in saliva (24%) (Supplementary table 1).

### Mosaic aneuploidy can originate in mitosis, meiosis I or meiosis II

The 33 observed duplications include 20 polysomies, 11 of which affect a whole chromosome and nine of which consist of the p-arm only (Supplementary table 1). Ten different chromosomes were affected by these polysomies (5, 8, 9, 12, 13, 14, 18, 21, 20, X). The origin of a trisomy can be determined by examination of the B-allele frequency pattern^6^. The absence of a third haplotype indicates that five of these events (three in chromosome 12 p-arm, one in chromosome 8 and one in chromosome X) have a mitotic origin. Eight events (one in chromosome 5 p-arm, one in each of chromosomes 9, 13, 14, 18 and 20, and two in chromosome 21) have a BAF pattern consistent with occurrence in meiosis I, where three haplotypes are observed near to the centromere. A pattern consistent with occurrence in meiosis II, with additional haplotypes present at the telomeres but not at the centromere, is observed in the remaining seven cases (three chromosome 12 p-arm, two chromosome 18 parm, one chromosome 13, and one chromosome 21) (Figure 3).

**Figure 3.**
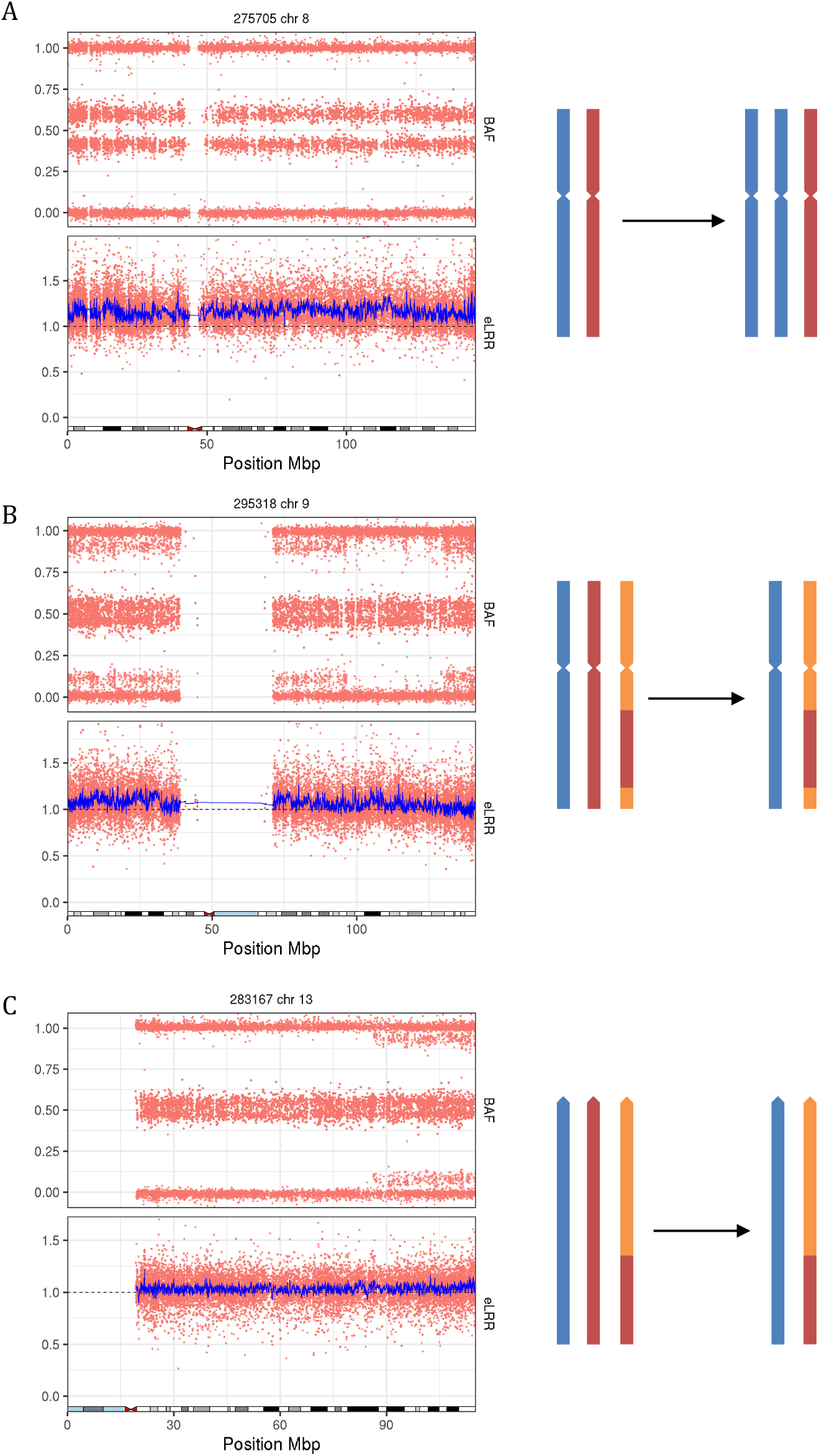
Origin of mosaic trisomies. A) A mosaic trisomy that has occurred during mitosis. B) A mosaic trisomy that has occurred during meiosis I. C) A mosaic trisomy that has occurred during meiosis II.

The 12 observed deletions include four mosaic monosomies, one in chromosome 7 and three in the X chromosome. One of the X chromosome monosomies has mosaic monosomy of the p-arm in 50% of cells and mosaic monosomy of the q-arm in 25 % of cells (Supplementary figure 1). In all of the observed monosomies, the B-allele frequency patterns are consistent with origination via mitotic nondisjunction, with two distinct haplotypes observed, rather than monosomy rescue^6^.

The 14 mosaic copy number neutral loss-of-heterozygosity events identified include five mosaic UPDs that comprise all or most of one arm of a chromosome (1q, 2p, 6p, 7q and 11q), four mosaic UPDs that affect an entire chromosome (three in chromosome 13 and one in chromosome 14) and five smaller loss-of-heterozygosity events. In one case, the UPD in the p-arm of chromosome 6 shows two different clonalities and is therefore likely to have arisen as two different events (Supplementary figure 2). Furthermore, we detected complex chromosomal events in several probands, including: deletion flanked by copy number neutral loss-of-heterozygosity spanning a total of 80.9Mb in chromosome 13 (Supplementary figure 3a); a duplication followed by UPD of the majority of chromosome 1 q-arm (Supplementary figure 3b); and a patient with a 59.9Mb duplication in chromosome 7, mosaic polysomy of the first 95 Mb of chromosome X and non-mosaic polysomy of the remainder of chromosome X (Supplementary figure 3c). We were unable to determine the origin of these events.

## DISCUSSION

We have identified MCAs using genotyping array data in 57/12,530 (0.45%) probands with severe developmental disorders. Fifty-four patients had single events, two had two independent events and one had 3 MCA events. Our findings are consistent with Sherman *et al*. in which 46 mosaic CNVs were identified in 12,077 probands with autism spectrum disorder (ASD)^27^. Following WES, only two (3.5%) patients with potentially clinically significant MCAs identified here have previously identified pathogenic or likely pathogenic SNVs, indels or CNVs. Compared with the cohort-wide diagnostic yield in the DDD study of ∼40%^28^, the observed enrichment of undiagnosed patients in this group suggests that most of these MCAs are diagnostic.

Clinical evaluation of the phenotypic and genomic data by an experienced clinical geneticist resulted in 44 diagnoses of MCAs that were either well-established pathogenic variants e.g., Mosaic tetrasomy 12p in Pallister-Killian syndrome, or where the chromosomal anomaly was classified as Pathogenic or Likely Pathogenic using the ACMG CNV classifier^29^ (Table 1). These comprised mosaic polysomies of chromosomes 12p, 18p and 20, mosaic duplications of chromosomes 5, 8, 11 and 17, mosaic deletions of chromosomes 2 and 22, mosaic loss-of-heterozygosity in chromosome 5 and a mosaic deletion-duplication-deletion in chromosome 18. Clinical features indicative of a mosaic event, including abnormalities of skin pigmentation, syndactyly and/or asymmetry, were observed in only seven of the 44 probands with a diagnostic finding. The remainder of the MCAs were interpreted to be variants of unknown significance.

MoChA is unable to distinguish between mosaic trisomies, mosaic tetrasomies or other mosaic polysomies. The six mosaic polysomies involving chromosome 12p are likely to be Pallister-Killian syndrome, where an isochromosome comprising two copies of chromosome 12p is present^30^. We also identify a case which is likely to be mosaic tetrasomy 5p. Only five cases of mosaic tetrasomy 5p, where an isochromosome consisting of two copies of the p arm of chromosome is present, have been reported to date^31^. In addition a case of an isochromosome consisting of two partial copies of 5p has been reported^32^. A small number of live-born cases of mosaic isochromosome 18p have been reported in the literature^33-35^, we identify a likely mosaic tetrasomy 18p.

Mosaic trisomy can occur by meiotic non-disjunction in the oocyte or sperm followed by trisomy rescue, or by mitotic nondisjunction at a later stage of development. Using genotyping array data, we were able to distinguish between mosaic polysomies occurring via nondisjunction at mitosis or meiosis, and 15/20 trisomies detected (75%) were meiotic in origin. The timing of the event has implications for counselling families, as some women have a higher rate of meiotic nondisjunction and therefore a greater recurrence risk^36,37^. This estimate is somewhat higher than Conlin *et al*., who found that 10/20 (50%) of trisomies had a meiotic origin^6^, and may reflect ascertainment differences between the cohorts.

The mosaic monosomies we detected all arose by mitotic nondisjunction, rather than monosomy rescue, which would result in homozygosity. This finding has important implications for recurrence risk, as for the former this is negligible whereas the latter raises the potential for gonadal mosaicism. We were unable to detect monosomy rescue as the method used is phase-based and therefore cannot detect events in runs of homozygosity^20^; however Conlin *et al*. also only reported mitotic events^6^. Similarly, our study can only detect mosaic UPD arising from trisomy rescue and resulting in heterodisomy as any events arising from monosomy rescue will result in isodisomy and lack heterozygous regions.

Two of the MCAs described here, a mosaic monosomy and a mosaic UPD, are in chromosome 7 and include the SAMD9 gene. In both patients pathogenic SAMD9 variants have previously been reported. Loss of chromosome 7 and UPD of 7q have previously been described in patients with MIRAGE syndrome, this is believed to be an adaptation to the growth-suppressing effect of the SAMD9 variants^38,39^.

We found MoChA to be a highly effective tool for detecting clinically relevant MCAs. Smaller subsets of the DDD cohort have previously been analysed for MCAs using alternative methods. Previously published analysis of structural mosaicism in SNP arrays from 1,303 DDD probands using MAD and triPOD described MCAs in 12 probands^3^.

However, neither MAD or triPOD detected all 12 of these events and it was shown that a combination of algorithms was necessary to maximise diagnostic yield. We tested 11 of these probands, and found nine of the previously reported events. MoChA identifies events found by MAD and missed by triPOD and vice versa. One of the events missing in our filtered MoChA dataset was a genome-wide paternal UPD. This event was found by MoChA, however the sample was removed by the MoChA default filters designed to exclude samples which are either contaminated or low quality DNA based on high phased BAF auto-correlation. The second event which is not found by MoChA was a UPD of chromosome 14 present in around two-thirds of cells; it is not clear why this event was not found by MoChA, however no mosaic UPDs with a cell fraction of >0.4 were detected. Additionally, a duplication in chromosome 17 not previously identified using MAD and triPOD was detected using MoChA. These results show that using more than one tool will increase the number of

MCAs detected. Furthermore, the previously published analysis of structural mosaicism in exome sequencing data from 4,911 DDD probands using MrMosaic described MCAs in nine probands, all of which were detected using MoChA^4^. In the same 4,911 probands an additional five events were detected using MoChA that were not detected using MrMosaic, including: two mosaic polysomies of chromosome 18p, one mosaic polysomy of chromosome 8, one mosaic UPD of chromosome 13 and one mosaic UPD of chromosome 2p.

Importantly, 23/26 (88.5%) of MCAs detected from saliva where blood was also available for testing could not be detected in blood-derived DNA. This result contrasts with our previous observation that mosaic *de novo* SNVs were observed at similar variant allele fractions in both blood and saliva^40^, and may suggest stronger negative selection against MCAs within blood lineages. One limitation of our study is that we only have data from two tissues, blood and saliva. While study of saliva yields more mosaic events than blood, mutations occurring later in embryonic development are likely to be present in a narrower range of tissues and we may therefore miss potentially diagnostic events by not having more tissue types available to study. Nonetheless, our observations highlight the importance of testing saliva (or other tissues) where possible to avoid missing mosaic structural events

There is currently a paucity of large-scale studies of MCAs in disease cohorts. Our results are comparable in both size and yield to those of Sherman *et al*., who report 46 mosaic CNVs in a cohort of 12,077 patients with ASD (0.38%)^27^, our MCAs included mosaic CNVs in 43 of our 12,530 patients (0.34%). Conlin *et al*.’s study of mosaic aneuploidies and UPDs in a cohort of 2019 patients referred to the Children’s Hospital of Philadelphia Clinical CytoGenomics laboratory had a higher yield than our study (30/2019, 1.5%)^6^. Our results add to this body of literature, but are likely to be an underestimate of the true diagnostic yield from MCAs in developmental disorders, due to under-ascertainment in the DDD study of cases who would have been previously diagnosed using prior clinical genetic testing (such as karyotyping and microarray analysis).

Our results show that rare mosaic chromosomal alterations are an important source of diagnoses in severe developmental disorders. The meiotic or mitotic origin of the mutation can often be determined through careful analysis of genotyping array data and has important implications for recurrence risk. This work suggests that routinely analysing SNP genotyping array data could provide potential diagnoses that are currently difficult to detect via WES, and that diagnostic yield will be increased by the analysis of saliva samples. We recommend that clinical teams consider the use of saliva-derived DNA for SNP array analysis for the investigation of neurodevelopmental disorders to complement genome-wide sequencing using blood-derived DNA.

## Data Availability

Diagnostic variants and phenotypes for probands included in this study are available via the DECIPHER database (https://deciphergenomics.org). Genotype array data is available in EGA.

## ACKNOWLEDGEMENTS

We thank the DDD participants and their families for their participation in this study. We thank Giulio Genovese for his assistance when running MoChA. We thank the Sanger Institute genotyping facility. The DDD study presents independent research commissioned by the Health Innovation Challenge Fund [grant number HICF-1009-003]. This study makes use of DECIPHER (http://decipher.genomics.org), which is funded by Wellcome. See Nature PMID: 25533962 or www.ddduk.org/access.html for full acknowledgement. This research was funded in part by the Wellcome grant [206194]. HF receives support from Wellcome grant 200990/A/16/Z. For the purpose of open access, the author has applied a CC BY public copyright licence to any Author Accepted Manuscript version arising from this submission.

## ETHICS DECLARATION

The study has UK Research Ethics Committee approval (10/H0305/83, granted by the Cambridge South Research Ethics Committee and GEN/284/12, granted by the Republic of Ireland Research Ethics Committee).

## AUTHOR CONTRIBUTIONS

Conceptualization: M.E.H.; Data curation: R.Y.E.; Formal analysis: R.Y.E., H.V.F.; Funding acquisition: M.E.H.; Investigation: R.Y.E.; Visualization: R.Y.E.; Writing-original draft: R.Y.E.; Writing-review & editing: R.Y.E., H.V.F., C.F.W., M.E.H., D.R.F.

---

**Supplementary figure 1.**
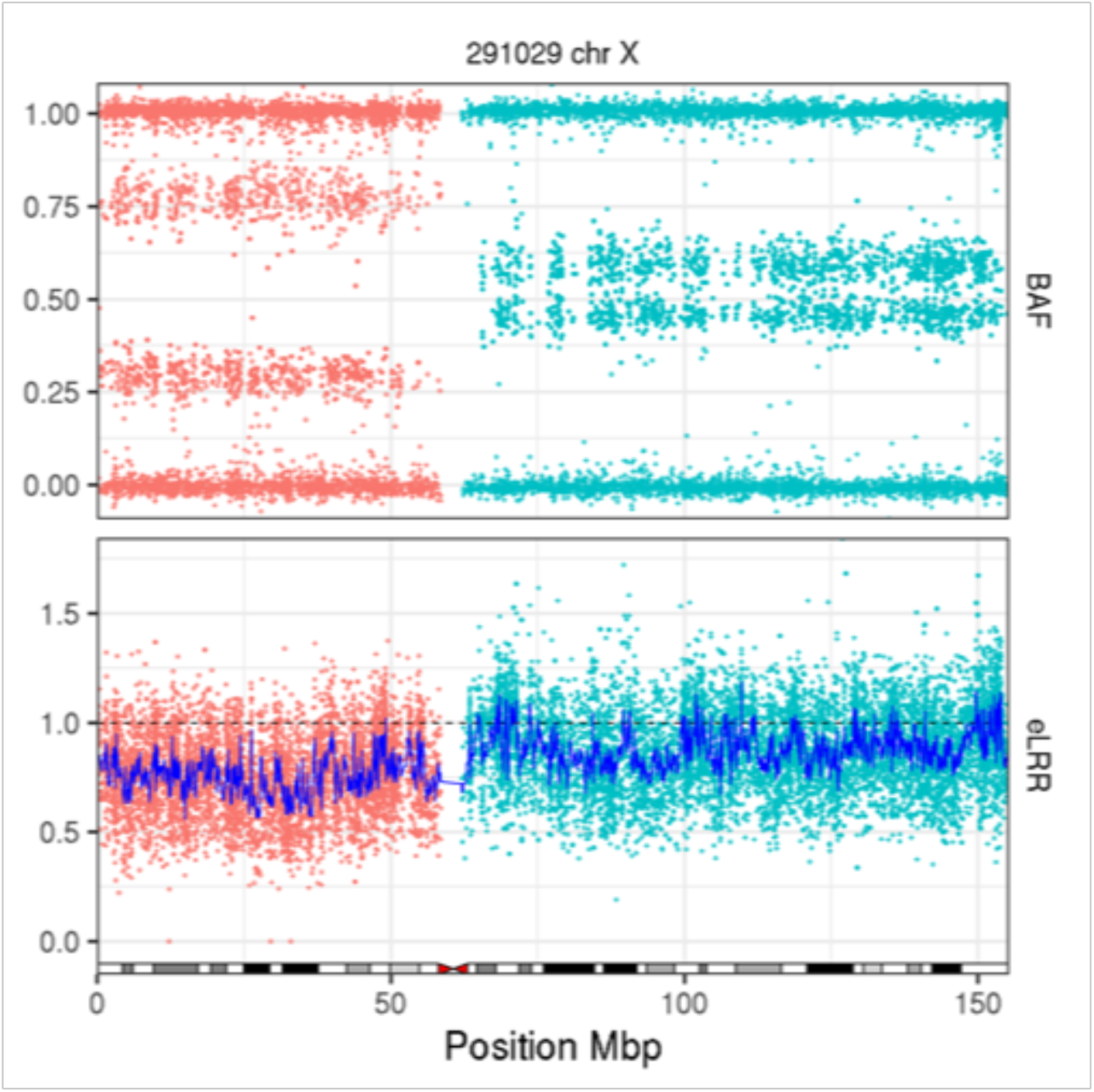
Mosaic deletion in chromosome X showing mosaic monosomy of the p-arm in 50% of cells (red) and mosaic monosomy of the q-arm in 25 % of cells (cyan).

**Supplementary figure 2.**
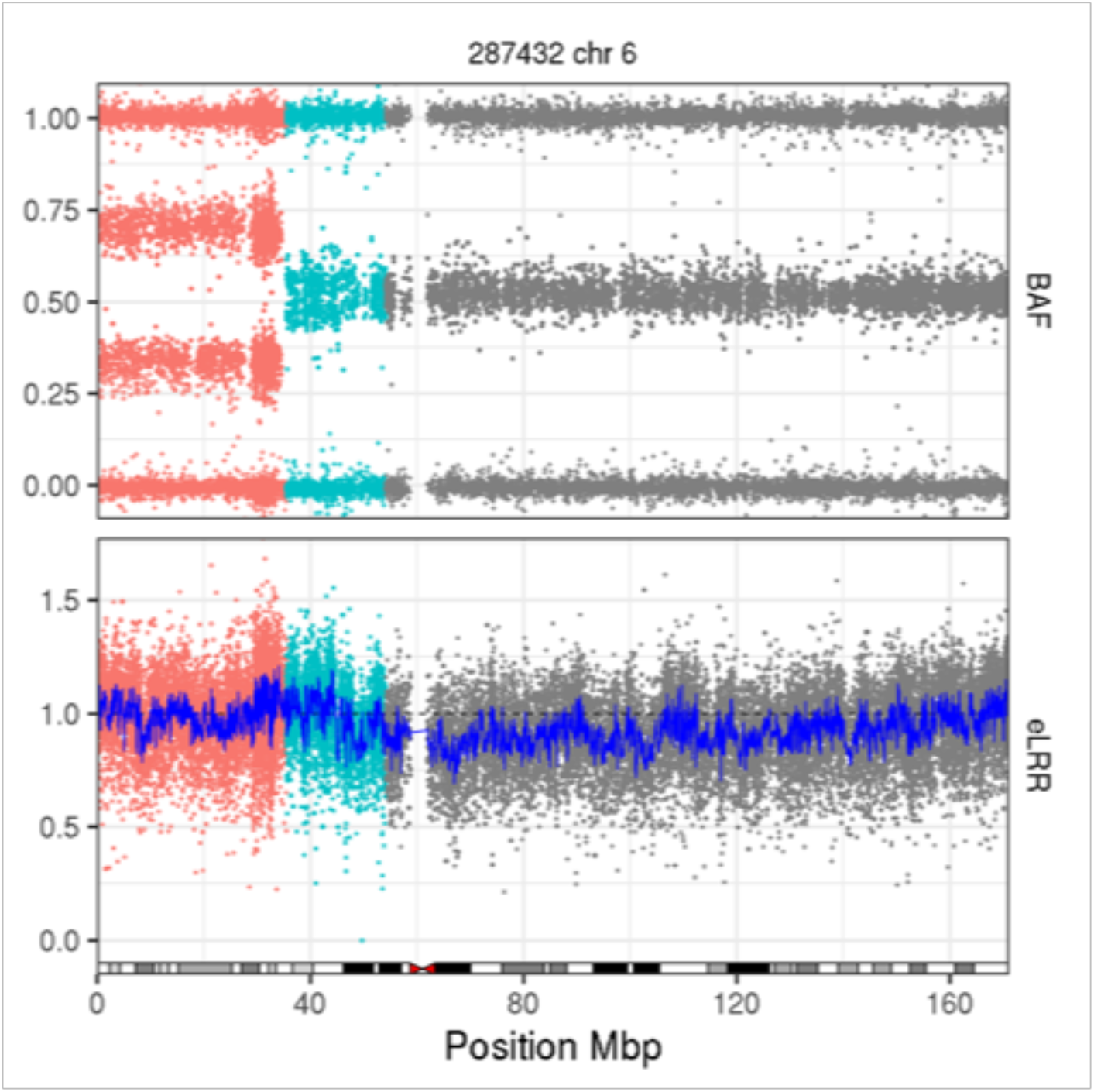
Mosaic UPD in the p-arm of chromosome 6 showing two different clonalities (red and cyan).

**Supplementary figure 3.**
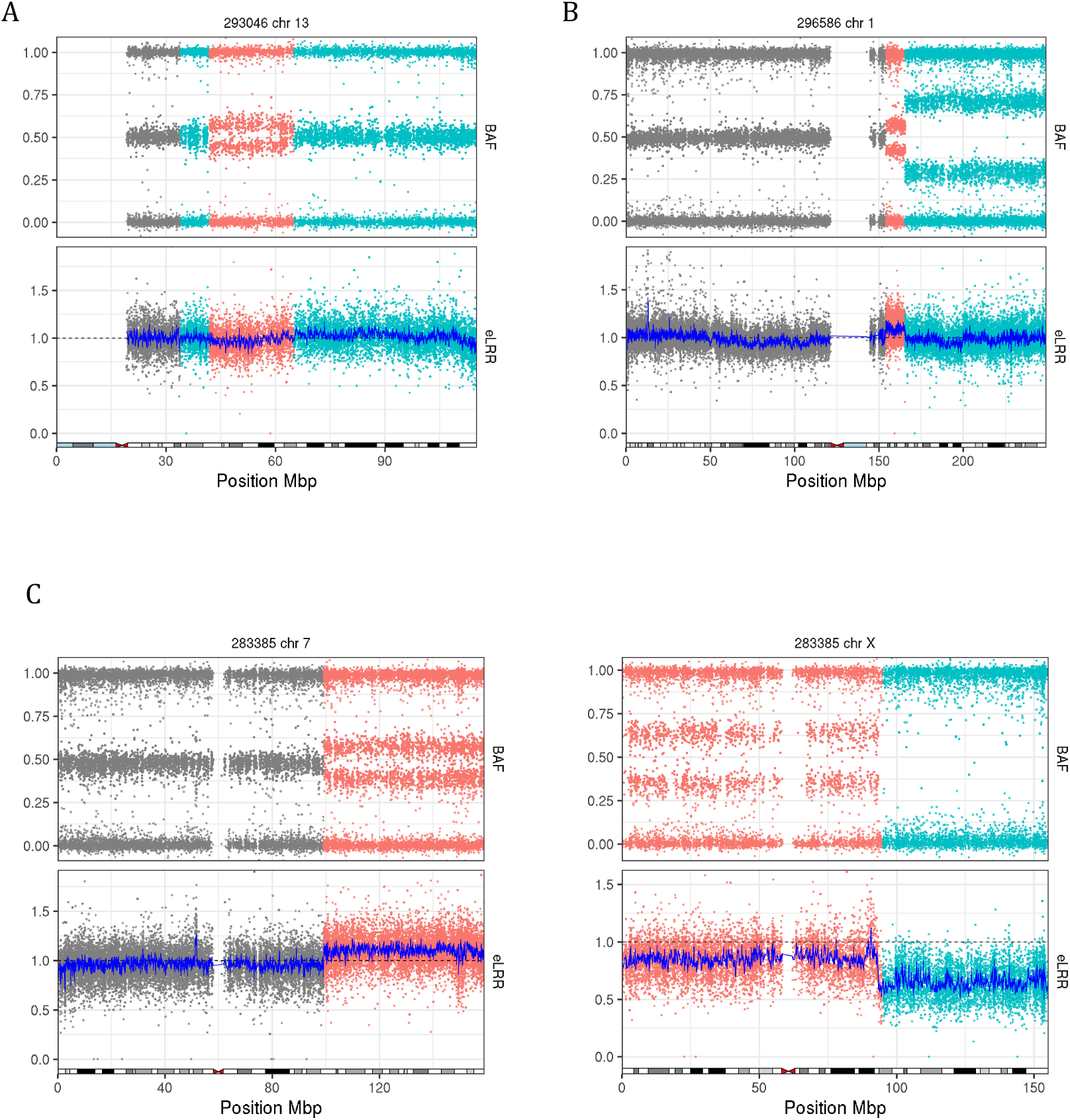
**Complex MCAs** A) A deletion (red) in chromosome 13 flanked by copy number neutral loss-of-heterozygosity (cyan). B) Chromosome 1 duplication (red) followed by UPD (cyan) of the majority of the q-arm. C) Duplication in chromosome 7 and polysomy in chromosome X comprising both mosaic (red) and non-mosaic (cyan) regions in the same patient. ^*a*^dup duplication, del deletion, upd uniparental disomy, upd (pat) paternal uniparental disomy ^*b*^w whole chromosome, p p-arm, q q-arm ^*c*^nd not done

